# Combining community wastewater genomic surveillance with state clinical surveillance: A framework for SARS-CoV-2 public health practice

**DOI:** 10.1101/2021.12.06.21267150

**Authors:** Ted Smith, Rochelle H. Holm, Ray Yeager, Joseph B. Moore, Eric C. Rouchka, Kevin J. Sokoloski, Daymond Talley, Vaneet Arora, Sarah Moyer, Aruni Bhatnagar

**Author notes:** Corresponding author: Ted Smith.

## Abstract

**Study objective:** To garner a framework for combining community wastewater surveillance with state clinical surveillance that influence confirmation of SARS-CoV-2 variants within the community, and recommend how the flow of such research evidence could be expanded and employed for public health response.

**Design, setting, and participants:** This work involved analyzing wastewater samples collected weekly from 17 geographically resolved locations in Louisville/Jefferson County, Kentucky from February 10 to November 29, 2021. Genomic surveillance and RT-qPCR platforms were used as screening to identify SARS-CoV-2 in wastewater, and state clinical surveillance was used for confirmation.

**Main results:** The results demonstrate increased epidemiological value of combining community wastewater genomic surveillance and RT-qPCR with conventional case auditing methods. The spatial scale and temporal frequency of wastewater sampling provides promising sensitivity and specificity to be useful to gain public health screening insights about community emergence, seeding, and spread.

**Conclusions:** Better national surveillance systems are needed for future pathogens and variants, and wastewater-based genomic surveillance represents opportune coupling. This paper presents current evidence that complementary wastewater and clinical testing is enhanced cost-effectively when linked; making a strong case for a joint public health framework. The findings suggest significant potential for rapid progress to be made in extending this work to consider pathogens of interest as a whole within wastewater, which could be examined in either a targeted fashion as we currently do with SARS-CoV-2 or in terms of a global monitoring of all pathogens found, and developing evidence based public health practice to best support community health.

**Thumbnail Box:** *What is already known on this subject?:* The primary approach for the genomic surveillance of SARS-CoV-2 relies on the sequencing of clinical COVID-19 samples. Variants of SARS-CoV-2 can also be tracked in community wastewater.

*What this study adds?:* We propose that, for comprehensive community surveillance, the first line of community pathogen screening should involve geographically-resolved wastewater samples collected at a regular frequency and employ both Next Generation Sequencing (NGS) and RT-qPCR. These results could then be compared with state clinical surveillance. This framework is a more comprehensive and cost-effective approach for surveillance in practice to catch community emergence, seeding, and spread.

*Policy implications:* Our results present a framework that could support the implementation of better surveillance policies directed to solve future community pathogen and variant detection. We anticipate this work can help public health officials implement rational community sampling schemes and develop sensible coordination with other clinical surveillance. The utility of this for COVID-19 extends to many other infectious disease models and other public health hazards such as toxic exposures.

## 1. Introduction

Many communities across the United States and world are using quantitative reverse transcription polymerase chain reaction (RT-qPCR) platforms to identify SARS-CoV-2 variants largely due to the low cost and effort involved. However, this approach has several distinct disadvantages. Most notably, there is often a lag in the availability of primers and probes once a variant of concern is declared, PCR cannot always distinguish between variants, and there is the loss of historical data about the wider range of mutations that may prove to be useful in post-hoc analysis (Boudet et al. 2021; Yaniv et al. 2021). Both the sequencing of clinical samples and community wastewater samples can further identify the circulating diversity of SARS-CoV-2 variants in an area (Izquierdo-Lara et al. 2020; Nemudryi et al. 2020; Fontenele et al. 2021; Jahn et al. 2021). As one of the communities in the United States employing next-generation sequencing (NGS) and comprehensive bioinformatic analysis of wastewater samples weekly for 9 months during the COVID-19 pandemic, Louisville /Jefferson County, Kentucky (USA) is uniquely positioned to provide insights and recommendations for this coupled approach and provide policy recommendations for other regions trying to implement genomic surveillance. To further leverage the public health response value of this approach, we have applied this genomic surveillance method with geographically-resolved wastewater sampling at 17 sites across Louisville/Jefferson County (Yeager et al. 2021). The aim of this study was to present the framework for combining community wastewater genomic surveillance with traditional clinical surveillance that influence confirmation of SARS-CoV-2 variants within the community, and to recommend how the flow of such research evidence could be expanded and employed for public health response.

## 2. Methods

### 2.1. Wastewater sample collection and handling

Wastewater was collected as a 24-hour time-weighted composite sample at 17 sites in Jefferson County, Kentucky, USA, from February 10 to November 29, 2021. Samples were collected once a week. The sites are a combination of street line utility holes and pump stations which are considered neighborhood or zip code level areas, and treatment centers covering larger geographic areas (Yeager et al. 2021). Samples were transported on ice to the University of Louisville for analysis by reverse transcription quantitative polymerase chain reaction (Rouchka et al. 2021).

### 2.2. Genomic surveillance

On a weekly basis, one sample for each site was submitted for sequencing, regardless of Ct value. As of week 40 (November 22, 2021 sampling date), a total of N=646 samples have been sequenced using NGS approaches. To enable high sensitivity, the samples were enriched for SARS-CoV-2 nucleic acids using the Swift Biosciences SNAP protocol for SARS-CoV-2. Samples were barcoded with the Swift Biosciences indexing kit and sequenced on an Illumina NextSeq 500 using the NextSeq Mid Output Kit v.5 at 300 cycles (2×150bp reads). The sequences were then directly aligned to the SARS-CoV-2 reference genome assembly (NC_045512.2) using BWA (Li 2013). Reads were then trimmed using Swift Biosciences primerclip (Addetia et al. 2020) to remove the primer read regions to avoid variant calling within these regions. The BCFTools mpileup (Li et al. 2009) utility was used to summarize the coverage of mapped reads and detected variants. Variants occurring with a frequency >= 0.05 with at least five sequences confirming the variant were filtered for further analysis. The pattern of mutations within each sample were there compared to variants of concern (VOC), variants of interest (VOI), and variants under monitoring (VUM). Those samples having at least 75% of mutations associated with a specific variant were then marked as likely containing that variant. Results of this analysis were available within the same week the sample was collected and were shared with public county and state public health officials.

### 2.3. Ethics

The University of Louisville Institutional Review Board classified this project as Non-Human Subjects Research (reference #: 717950).

## 3. Results and Discussion

The University of Louisville (UofL) has led the application of wastewater-based epidemiology (WBE) as a partner to both Louisville Metro Public Health and Wellness (LMPHW) and to the Kentucky Department of Public Health (DPH), beginning in May 2020. The initial focus of these arrangements was on quantifying virus recovered in community and correctional facility wastewater to help inform public health officials about intensity and spread of infection. However, by February 2021, the work expanded to include full NGS at the UofL of the recovered viral RNA fragments to arrive at probabilistic estimates of the likelihood of the presence of variants of SARS-CoV-2 in community catchment areas across the Louisville/Jefferson County, Kentucky area. The work included Centers for Disease Control and Prevention (CDC) variants of concern (VOC) and variants of interest (VOI) as well as some variants of attention but with no classification. The initial surveillance starting in February 2021 included: B.1.1.7, B.1.351, B.1.427, B.1.429, P.1, R.1, B.1.618, B.1.623, B.1.617.1, B.1.617.2, B.1.617.3, and B.1.621. On July 5, 2021, we added C.37. On July 26, 2021, we added B.1.525, B.1.526, and P.2. The Omicron (B.1.1.529) variant was added on November 30, 2021. Genomic sequencing results were typically available 4 days after wastewater sample collection. During the initial four months of genomic analysis of the wastewater sampling, there were several opportunities to explore the utility of this type of sampling at the community and regional level. What emerged was a loose coupling between the wastewater analysis effort and the state’s clinical genomic surveillance. Collaboration between state public health and academic centers of excellence, formal or informal, presents an excellent opportunity for more complete situational awareness and shared learning. Initiation of these collaborations often only involve reaching out to the counterparts in the academic center or the state public health and sharing ideas and needs. We saw this simple exchange of ideas between UofL and DPH develop into the program that has produced successful scenarios, some of which we present here.

We discovered three scenarios that demonstrated complementary data from each activity. First, because community wastewater sampling is a form of pooled testing, it effectively samples 100% of the population connected to the wastewater system. Thus, it is far more likely to detect an emerging variant than would be found through sparse sequencing of clinical nasopharyngeal swabs. This was demonstrated in early April 2021 when our wastewater genomic surveillance identified the Gamma (P.1) SARS-CoV-2 variant and UofL communicated that to the county and state for geographically and demographically targeted public health response. The variant was confirmed one week later when the first clinical case of that variant within the same wastewater catchment area was identified. Since then, there have been numerous comparisons between the catchment areas and clinical trends with this same “Wastewater Community Screen and Clinical Confirm” approach. Our observation is similar to Jahn et al. (2021), who reported detection of B.1.1.7 in wastewater treatment centers samples in mid-December, and only later were these reported in clinically derived community data.

The second scenario occurred in late April 2021 when the DPH lab sequenced the clinical samples from a rural senior living center outbreak reported to be caused by the B.1.1.316.1 (R.1) variant. The DPH requested a check with the UofL wastewater database to see if that particular variant had been detected at any time anywhere in the county to better understand spread risk while the United States government SARS-CoV-2 Interagency Group was making a determination of its classification (Cavanaugh et al. 2021). Fortunately, there was no evidence that R.1 had spread to Louisville/Jefferson County.

In the third instance, the B.1.526 variant was detected in one of the collection sites on August 26, 2021, a time at which Delta was dominate across all locations. Subsequent clinical sequencing yielded a B.1.526 sample collected on September 17, 2021 from an adjacent zip code (Supplement Figure A1). This illustrates the early detection capabilities of the wastewater surveillance.

Since February 2021, weekly updates from UofL as academic partners are provided to LMPHW, graphically representing the presence of wastewater variants within a rolling 4-week period (Supplement Figure A2). On a weekly basis, it was not unusual for variants to appear or disappear, and a moving 4-week period allowed stability as to the scale of variant presence.

Moving reporting from a formal technical document to less than a dozen slides allowed rapid understanding even among non-genomics and non-bioinformatics professionals and has been an accepted and efficacious data deliverable format as received by LMPHW. While other genomic work has focused on treatment centers wastewater samples (Nemudryi et al. 2020; Crits-Christoph et al. 2021; Smyth et al. 2021), the benefit of our geographically-resolved sampling design is that it allowed geographically-precise identification of variant emergence and neighborhood spread on a weekly basis. Our results noted variants observed in smaller wastewater catchment areas, such as neighborhood street line utility holes, were not uniformly observed downstream at the associated treatment centers. Other genomic wastewater SARS-CoV-2 sequencing work in the United States has also showed this similar utility of regular temporal data (Nemudryi et al. 2020; Crits-Christoph et al. 2021; Fontenele et al. 2021; Smyth et al. 2021).

Our work leveraged a foundational collaboration that existed prior to the pandemic between the UofL, LMPHW, and the DPH, and the Louisville Metropolitan Sewer District. Our genomics and bioinformatics laboratories have been involved in whole genome metagenomics in other contexts, typically metagenomics populations within individuals. Financially, significant components of the sampling and viral analysis were paid for through other mechanisms including the American Rescue Plan and related federal public health funds. Additionally, UofL is also concurrently funded by the NIH through Institutional Development Award (IDeA) program, by the CDC for the wastewater sampling and testing, and by the Commonwealth of Kentucky for wastewater monitoring of correctional facilities. These relationships provided the project with strong communication channels with our state public health commissioner and the CDC National Wastewater Surveillance System (NWSS) leadership that have enabled translation from academic lab settings to sustainable ongoing deployment of the public health response developed in this framework.

This work has helped answer important questions many communities in the United States and around the world have about the utility of conducting wastewater surveillance as part of genomic surveillance for infectious diseases. Over the course of the pandemic, one of the greatest challenges has been the need to span fields of expertise and capability to arrive at useful solutions. It is already showing some promise in Louisville/Jefferson County and our results inform the roadmap for global adoption and long-term general capability. For example, much of the data from the wastewater field has relied on PCR analysis of wastewater samples gathered from wastewater centers or from college dormitories. Our framework (Figure 1) provides valuable insights for other communities about the feasibility and value of NGS analysis for SARS-CoV-2 variants in sampling scenarios that utilize smaller catchment areas. We anticipate this work will help public health officials implement improved community sampling schemes and develop coordination with other clinical surveillance. The utility of this approach for COVID-19 certainly extends to many other infectious disease models and other public health hazards such as toxic exposures.

**Figure 1.**
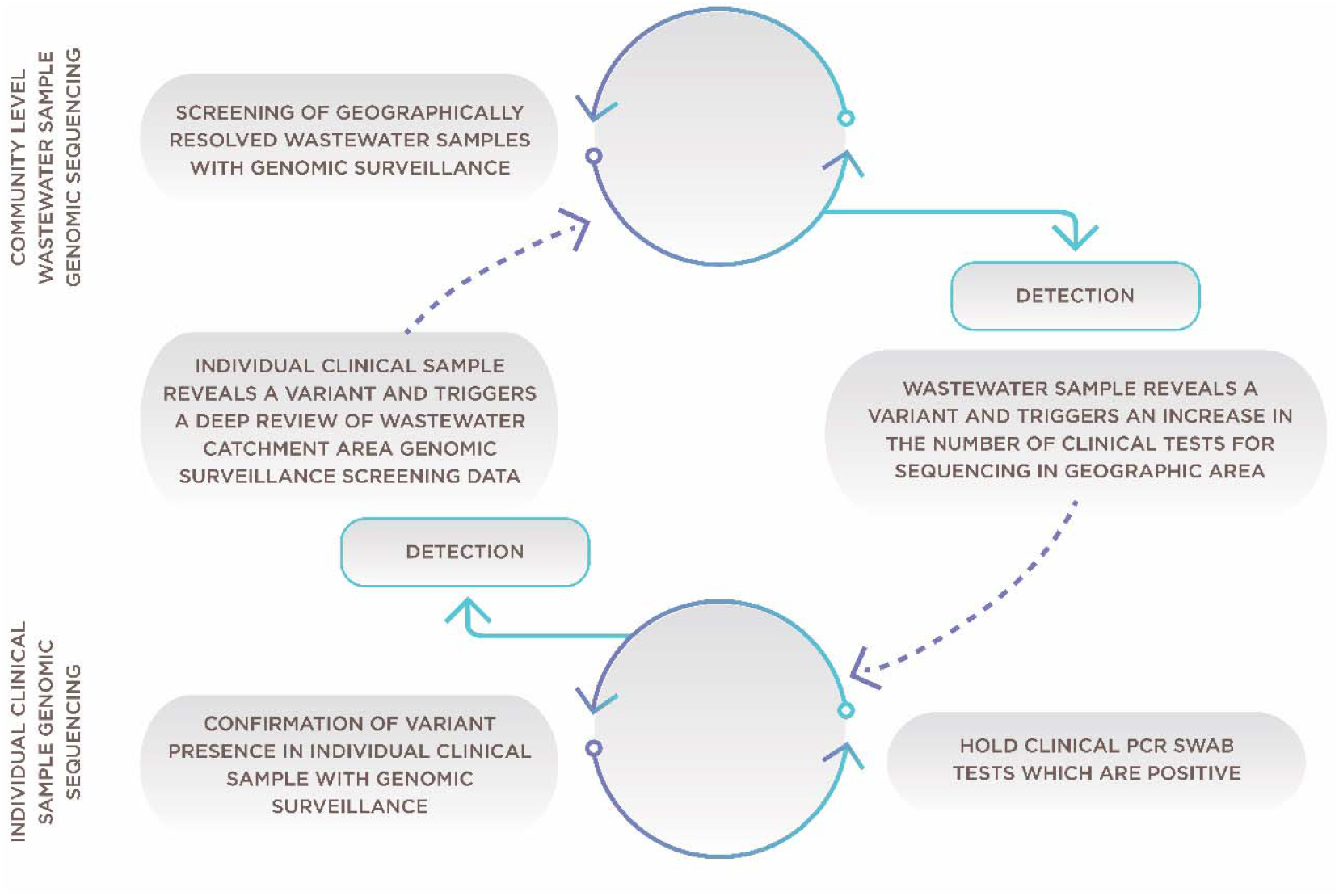
Framework for combining community wastewater genomic surveillance with state clinical surveillance

## 4. Future Policy

This research has identified six key policy areas for exploration. These include:

1. The spatial scale of wastewater sampling that provides optimal sensitivity and specificity to gain insights for public health response. For example, it may be less useful to know a variant is present in a wastewater catchment area covering half the population than to have neighborhood-scale data. Conversely, there is likely quickly diminishing value in very small areas when cost and effort are considered.
2. The temporal frequency of community wastewater samples providing an optimal balance of “early warning” and also information about seeding and spread. Some of this issue has been addressed in the analysis of viral concentrations in community sampling. However, the importance of identifying emerging variants that escape vaccination creates a case for getting ahead of spread over time.
3. The coupling with clinical genomic surveillance has been anecdotally substantiated but would be more useful in future situations and in other communities if there were a systematic analysis of the relationship between spatiotemporal coverage of clinical genomic surveillance and of wastewater genomic surveillance. For example, it may be possible to derive a useful level of clinical testing as a function of the wastewater catchment area where a variant is first discovered. Conversely, a clinical case with a new variant first identified at a location could trigger a series of nested catchment area samples to be collected which is one of the classic approaches in “sewer tracing” dating back to cholera.
4. Community seeding and spread of different variants is an area of analysis that could improve understanding of possible entry points in communities and patterns of spread. For example, initial analysis of historic data in Louisville/Jefferson County suggests neighborhoods near our air logistics hub appear to be more likely to initially establish new variants over time. It is possible our data could be connected to the larger database for Kentucky and perhaps the country to provide insights into characteristics of places that could inform new public health interventions.
5. The role vaccination plays in suppressing spread of variants is difficult to discern from clinical testing alone. We propose wastewater samples to estimate levels of suppression associated with vaccination coverage in an area to inform transmission rates in communities of high and low vaccination coverage.
6. Determining at a high level what the cost-benefit tradeoffs are for different configurations of wastewater and clinical genomics surveillance can be established using Louisville/Jefferson County’s historic costs for both wastewater genomics and clinical genomic testing.

## 5. Limitations

Wastewater samples may lack the RNA quality for longer sequence reads necessary to identify multiple mutations in a single RNA amplicon. Wastewater detection relies on the capacity of a pathogen to be shed into the wastewater, and pathogens or variants with altered shedding may create new challenges for detection. Wastewater surveillance cannot provide data in residential areas that are not served by municipal sewer systems and thus excludes some residences even in an urban area such as Jefferson County. The limit of detection for wastewater surveillance (i.e., the smallest number of infections in a community that can be detected in wastewater) is not well established, thus wastewater surveillance cannot be used to determine whether a community is actually free from SARS-CoV-2 infections and should be used in combination with clinical results.

## 6. Conclusion

This is the ideal time to improve public health surveillance system for pathogens, and wastewater-based genomic surveillance represents a feasible and comprehensive complement to established approaches. Further, the addition of genomic sequencing into the wastewater surveillance framework may provide a more comprehensive assessment of regional viral infection surveillance when compared to clinical testing alone. The value of this project to the public health and clinical scientific community rests in our ability to produce compelling evidence that COVID-19 genomic surveillance is cost-effectively enhanced through complementary wastewater and clinical testing. There is important value in using a broad perspective approach to identifying low abundance variants early from a few samples. A natural extension of this work is in considering pathogens of interest as a whole within wastewater, which could be examined in either a targeted fashion as we currently do with SARS-CoV-2 or in terms of a global monitoring of all pathogens found, to develop evidence based public health practice to best support community health.

## Data Availability

All data produced in the present study are available upon reasonable request to the authors.

## Supplement A

**Figure A1.**
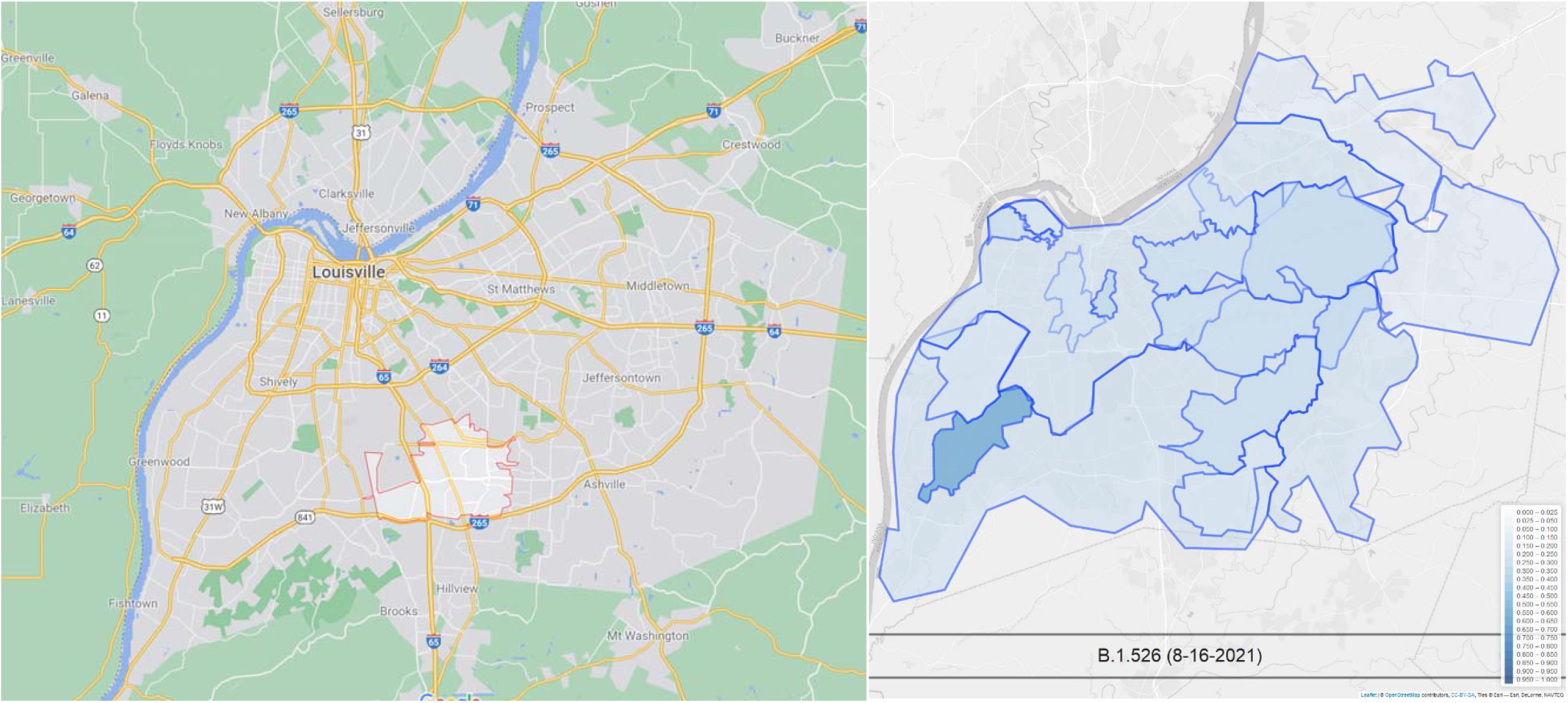
Clinical sequencing yielded a B.1.526 sample from an individual in the 40219 zip code (left) from a sample on 9/7/2021. Examination of the wastewater samples indicated the presence of B.1.526 from a nearby wastewater collection site two week prior, on 8/16/2021.

**Figure A2.**
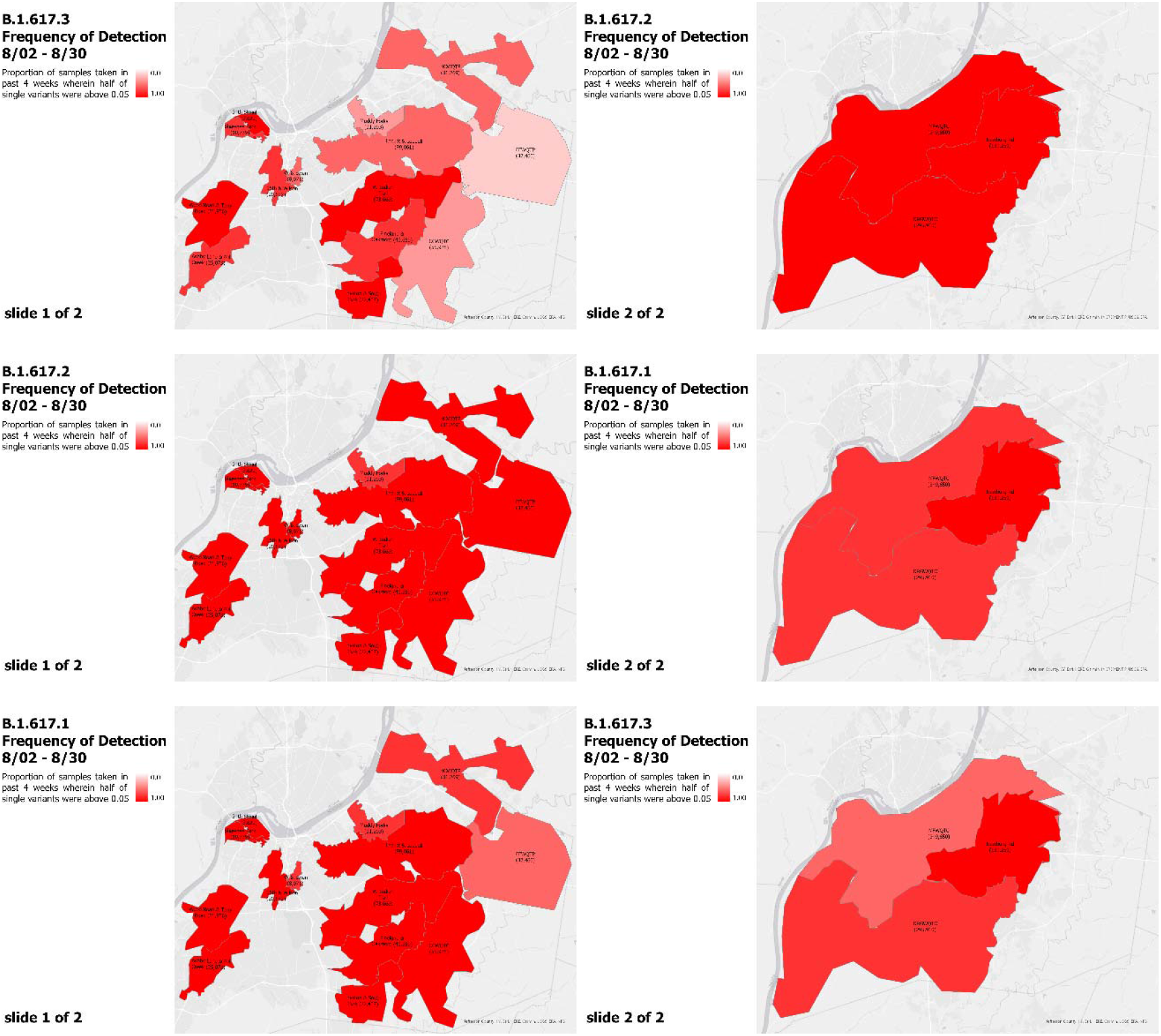
August 2021, genomic surveillance of SARS-CoV-2 in wastewater data in Louisville /Jefferson County, Kentucky as presented weekly by academic partners to Louisville Metro Health and Wellness.

